# Diabetes in African Youth, Improving Glucose Time-In-Range (DAYTime): A Randomized Controlled Trial

**DOI:** 10.1101/2023.03.19.23287358

**Authors:** Thereza Piloya-Were, Catherine Nyangabyaki, Elizabeth Pappenfus, Muna Sunni, Brandon M. Nathan, Expeditus Ahimbisibwe, Ezrah Trevor Rwakinanga, Lin Zhang, Silver Bahendeka, Antoinette Moran

**Author notes:** <u>Corresponding Author</u> Thereza Piloya-Were, Department of Paediatrics, College of Health Sciences., Makerere University, Kampala, Uganda.

## Abstract

**Introduction:** Disparities in healthcare access and clinical outcomes exist in individuals with type 1 diabetes (T1D) in less- versus well-resourced countries. An observational study of East African youth with T1D found an average haemoglobin A1c level of 11%. Blinded continuous glucose monitoring (CGM) demonstrated extremes of both hyper- and hypoglycemia. This randomised clinical trial (RCT) aims to test the hypothesis that enabling Ugandan youth with T1D to monitor glucose levels with CGM technology will improve glucose time-in-range (TIR, glucose levels 3.9-10.0 mmol/L).

**Methods and Analysis:** Ugandan youth with T1D (n=180, age 4-26 years) will be randomized over five years, August 15, 2022-August 14, 2027, at Mulago or Nsambya Hospitals, Kampala. Half will be placed on <u>unblinded</u> Freestyle Libre 2 CGM for 12 months. The other half will be given sufficient test strips for 3x daily self-monitoring of blood glucose (SMBG) while wearing <u>blinded</u> CGM for 6 months (control group). Current standard-of-care is 2-3 test strips per day. The control group will switch to unblinded CGM months 7-12. All subjects will receive monthly diabetes education. The primary endpoints are 1) the 6-month change from baseline in glucose TIR in those wearing the unblinded CGM compared to those performing SMBG, and 2) cost analysis of CGM compared to 3x/day SMBG, to determine whether this technology is cost effective in a less-resourced country. The primary hypotheses will be tested by linear mixed effects models.

**Ethics and Dissemination:** The protocol was approved by the Mulago Hospital Research Ethical Committee (MHREC 2173), Uganda National Council of Science and Technology (HS2129ES), and the University of Minnesota IRB (STUDY00013430). Results will be disseminated at scientific meetings, policy briefings at the Ugandan Ministry of Health, and peer reviewed journal publications.

**Clinicaltrials.gov registration:** NCT05454176

**Protocol Version** 4.0, December 5, 2022

**Strengths and Limitations of this Study:**

- The study will be conducted at the two largest diabetes clinics in Uganda, involving a large number and wide age range of children and young adults from a less-resourced nation.
- All participants will receive extensive diabetes education.
- The University of Minnesota will monitor the study and will provide ongoing research education, training the Ugandan team for involvement in this and future RCTs.
- The inclusion of Ministry of Health economists allows assessment of the real-world practicality of this intervention.
- The study is being conducted in an urban setting, with no rural comparison.

## INTRODUCTION

In 2017, the International Diabetes Federation (IDF) reported ∼1.1 million children living with type 1 diabetes (T1D) world-wide. Of these, 50,600 were known to be living in Africa, with ∼18,300 new cases diagnosed each year (1). The prevalence continues to rise as detection, reporting, and survival improve (2). In less-resourced regions, even minimal diabetes care is beyond the financial means of many families (3). However, in many countries, the organizations CDiC® (Changing Diabetes in Children, Novo Nordisk) and LFAC (Life for a Child), working with national ministries of health, have helped fill that gap by training healthcare teams and providing insulin, test strips and patient education materials. In Uganda, approximately 1500 children and adolescents with T1D are registered under CDiC® care, and 312 with LFAC.

Even with these major improvements in healthcare delivery, achieving metabolic control has been difficult. A recent study performed in Uganda and Kenya found very poor control, with extreme levels of both hypo- and hyperglycemia in children and youth followed monthly in dedicated diabetes clinics with trained personnel, receiving adequate quantities of human insulin, diabetes education, and sufficient test strips to measure glucose levels 2-3x per day (4). Using a blinded continuous glucose monitor (CGM), this observational study found a mean glucose level of 231±86 mg/dl (12.8±4.8 mmol/L), with a coefficient of variation of 48±21%. Only 30±19% of time was spent in the target glucose range of 70-180 mg/dl (3.9-10.0 mmol/L), 13±16% of time the glucose was <70 mg/dl (3.9 mmol/L), and 7±8% of time the glucose level was <55 mg/dl (3.1 mmol/L). Very low hypoglycemia (glucose <55 mg/dl, 3.1 mmol/L) occurred in 81% of participants, averaging 5 events per week with an average duration of 140±79 minutes per event. Despite this degree of hypoglycemia, haemoglobin A1c (HbA1c) levels averaged about 11%. In contrast, average HbA1c levels in American youth average 8.1-9.3% (5, 6).

US T1D Exchange data show that more frequent glucose monitoring is associated with lower HbA1c levels in youth (7). Self-monitoring of blood glucose (SMBG) is an essential but expensive component of diabetes care. International paediatric guidelines recommend SMBG 6- 10 times per day (8). The American Diabetes Association (ADA) recommends SMBG at least 4 times per day (before meals and at bedtime), plus additional testing throughout physical exercise, during hypoglycemia and hyperglycemia and before driving, all of which can require at least 6- 10 tests per day (9). East African youth are rarely able to test more than 2-3 times per day (4).

According to one international report (10), the level of care currently available in Uganda is considered to be within the “intermediate care” tier. While this represents an improvement compared to a decade ago, high HbA1c levels and the high percentage of time spent in both hypo- and hyperglycemia place these youth at significant risk for short- and long-term diabetes complications and thus current methods of management are inadequate.

CGM is rapidly becoming standard-of care in well-resourced nations, but is virtually unknown in East Africa due to cost and lack of availability. It has been shown to decrease HbA1c levels, increase glucose time-in-range, and reduce hypoglycemia (11-16). In addition, parents of young children report increased peace of mind with CGM technology (16). The ADA recommends that access to CGM devices should be considered from the outset of a diagnosis of T1D (9).

CGM devices are currently too expensive to consider in less-resourced settings, but as they become more ubiquitous worldwide, prices are coming down. In the UK, a 2018 report suggested that flash CGM monitoring could be cost saving for people testing blood glucose levels 6 or more times per day (17). A 2020 US cost comparison found that flash glucose monitoring was equivalent in price to 3x/day SMBG (18). Avoiding diabetic ketoacidosis (DKA) and severe hypoglycemia, preventing long term micro- and macrovascular complications, providing the opportunity to achieve normal life milestones (education, sports participation, employment, marriage, having children), and being able to expect normal longevity all need to be figured into cost-benefit calculations.

If CGM leads to a significant improvement in diabetes metabolic control in Ugandan youth by reducing hypo-and hyperglycemia, then the ethical question is not whether to make it available to patients in less-resourced settings, but how to make it affordable. This is similar to the issues that arose when HIV/AIDS therapies first became available. Such discussions and decisions must be guided by data obtained in the unique settings found in less-resourced countries, and must include cost-benefit analyses. This protocol aims to obtain these data.

## METHODS AND ANALYSIS

The protocol is reported following the Standard Protocol Items Recommendations for Interventional Trials (SPIRIT). The clinical trial is registered with clinicaltrials.gov (NCT05454176).

### Overall Goal and Hypotheses

The overall goal is to determine whether CGM use in Ugandan children and young adults with T1D will improve overall diabetes control as determined by increased glucose time-in-range. We hypothesize that 6 months of CGM use will lead to a significant increase in glucose TIR compared to 3x/day SMBG. We further hypothesize that these improvements will be cost-effective.

The study provides an opportunity for the experienced Minnesota research team to train and mentor Ugandan paediatric diabetes teams in the conduct of randomized controlled clinical trials (RCTs), thus increasing local research capacity. The Ugandan sites have experience conducting observational studies, but minimal experience with RCTs.

### Objectives

The primary objectives of this study are:

1. To determine if patients’ ability to continuously observe interstitial glucose levels for 6 months using the Freestyle Libre 2 CGM device improves glucose TIR from baseline assessment. The change in glucose TIR while wearing the unblinded CGM will be compared to change in TIR in patients performing 3x/day SMBG (wearing a blinded CGM for endpoint measurement).
2. To perform a cost analysis on flash glucose monitoring compared to 3x/day SMBG, and to determine whether this technology is cost effective in the setting of a less-resourced nation.

Secondary Objectives are to assess the change-from-baseline impact of unblinded CGM on:

1. Percent TIR at 12 months
2. Percent time with glucose 180-250, >250, <70, and <54 mg/dl (10.0-13.9, >13.9, <3.9, and <3.0 mmol/L) at 6 and 12 months
3. HbA1c at 6 and 12 months
4. Patient satisfaction and quality of life at 6 and 12 months
5. Glucose variability (coefficient of variation, CV) at 6 and 12 months

There are also training objectives for the Ugandan research teams and the Kampala diabetes community.

### Study Design

This is a randomized, non-blinded, phase 4 clinical trial. Subjects will be randomly assigned 1:1 to the CGM Group or the Control Group. Randomization is stratified by clinic and by age group (4-11, 12-18, and 19-26 years) with approximately equal numbers in each age group and clinic location. At baseline, a 1-2 week initial assessment will be performed where the ability to wear and return the sensor can be demonstrated as an entry criterion for study randomization. After this, 180 children and young adults with T1D will be randomized into the 12-month clinical trial in four annual cohorts of about 45 patients each.

The study design is shown in the Figure. Half of subjects (n=90, “CGM Group”) will receive unblinded FreeStyle Libre 2 CGM for the entire 12 months. Half of subjects (n=90, “Control Group”) will be given sufficient test strips for 3x daily SMBG levels months 1-6, and they will wear blinded CGM for endpoint assessment. Months 7-12 they will switch to unblinded CGM.

**Figure:**
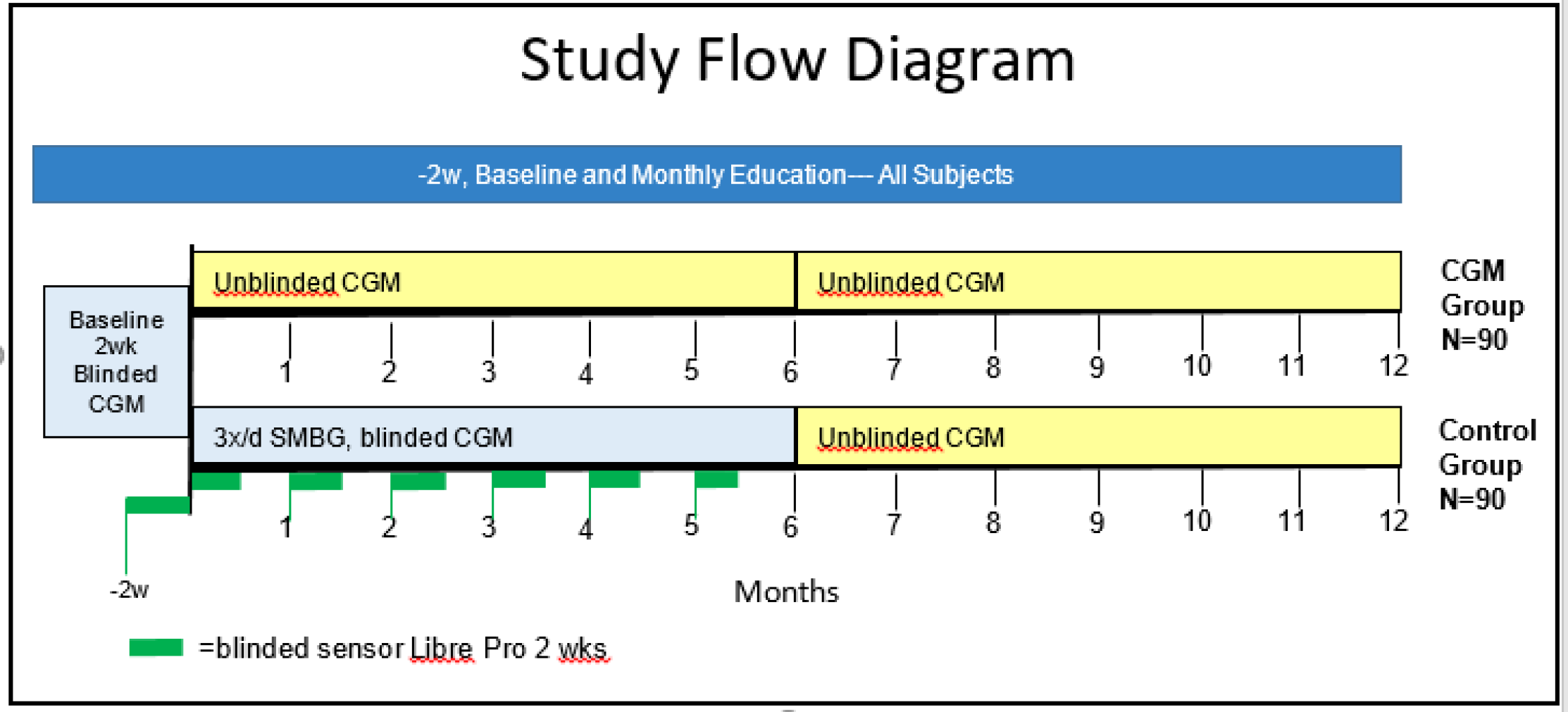
Study Flow Diagram.

Primary endpoint assessment will occur after 6 months of unblinded CGM use (months 1-6 for the CGM Group and months 6-12 for the Control Group). The first 6 months of the Control Group (3x/day SMBG while wearing blinded CGM) will serve as a control. The CGM Group will receive an additional 6 months (months 7-12) of unblinded CGM therapy to determine the effects of 1 year of CGM. The Table lists the schedule of study assessments.

**Table.** Overview of the patient schedule of activities.

**Table. Overview of the patient schedule of activities**
| Time (-2 weeks, then month) | -2wk | 0 | 1 | 2 | 3 | 4 | 5 | 6 | 7 | 8 | 9 | 10 | 11 | 12 |
| --- | --- | --- | --- | --- | --- | --- | --- | --- | --- | --- | --- | --- | --- | --- |
| Visit Number | 1 | 2 | 3 | 4 | 5 | 6 | 7 | 8 | 9 | 10 | 11 | 12 | 13 | 14 |
| Informed Consent/Assent | X |  |  |  |  |  |  |  |  |  |  |  |  |  |
| Initial Eligibility Screening | X |  |  |  |  |  |  |  |  |  |  |  |  |  |
| Final Eligibility, Randomization |  | X |  |  |  |  |  |  |  |  |  |  |  |  |
| Complete Medical History | X |  |  |  |  |  |  |  |  |  |  |  |  |  |
| Interim Medical History |  | X | X | X | X | X | X | X | X | X | X | X | X | X |
| Physical Activity History |  | X | X | X | X | X | X | X | X | X | X | X | X | X |
| Pregnancy inquiry (as appropriate) | X | X | X | X | X | X | X | X | X | X | X | X | X | X |
| Prior/Concomitant Meds | X | X | X | X | X | X | X | X | X | X | X | X | X | X |
| Adverse Event Assessments | X | X | X | X | X | X | X | X | X | X | X | X | X | X |
| Daily Insulin U/kg/day, Type of Insulin | X | X | X | X | X | X | X | X | X | X | X | X | X | X |
| Severe Hypoglycemia History | X | X | X | X | X | X | X | X | X | X | X | X | X | X |
| Physical Exam | X |  |  |  |  |  |  |  |  |  |  |  |  |  |
| Limited/Directed Physical Exam* |  | X | X | X | X | X | X | X | X | X | X | X | X | X |
| Clinic POC HbA1c | X |  |  |  | X |  |  | X |  |  | X |  |  | X |
| CGM Sensor Placement** | X | X | X | X | X | X | X | X | X | X | X | X | X |  |
| CGM and/or Meter Download |  | X | X | X | X | X | X | X | X | X | X | X | X | X |
| Questionnaires | X |  |  |  |  |  |  | X |  |  |  |  |  | X |
| Insulin, Test Strips Dispensed |  | X | X | X | X | X | X | X | X | X | X | X | X | X |
| CGM Dispensed | X | X | X | X | X | X | X | X | X | X | X | X | X |  |
| Patient Education | X | X | X | X | X | X | X | X | X | X | X | X | X | X |

\*Limited exam = pulse, BP, height, weight, injection/insertion sites, additional exam for cause as determined by interim history
\*\*Participants will also be taught to insert the sensors themselves at home if insertion times don't coincide with clinic visits.

### Study Setting, Recruitment and Consent

This study will be conducted in two urban T1D clinics in Kampala, Uganda, at Mulago and St. Francis, Nsambya Hospitals. Patients will be recruited by their paediatricians, who are investigators in this study, through their paediatric diabetes clinics. The Mulago T1D clinic is located at the Mulago National Referral Hospital, a teaching hospital for Makerere University. T1D care is supported by a paediatric endocrinologist, residents and 2 nurses. The Nsambya T1D clinic is located St. Francis Hospital, Nsambya, which is a teaching hospital for Mother Kevin Post Graduate Medical School, Uganda Martyrs University. The diabetes team is headed by a paediatrician and is supported by 2-3 nurses plus residents. Each clinic follows about 300 children and young adults age 0-28 years. St. Francis Hospital, Nsambya is a private not-for-profit missionary hospital while Mulago is a public government hospital. The patients seen at St. Francis Hospital, Nsambya have a moderately higher socioeconomic status than those seen at Mulago hospital, but only a few are able to afford better health care or insurance. Both clinics are supported by the CDiC® program, with the children receiving free insulin (Regular as Actrapid and Neutral Protamine Hagedorn as Insulatard), 2-3 glucose strips per day for SMBG, measurement of HbA1c levels every three months, diabetes education, and occasionally an annual diabetes camp for psychosocial support. CGM use is rare in Uganda; we will not enroll any participants with current CGM use.

Written informed consent will be obtained from all participants. Parents/guardians will provide consent for children age <18 years, youth ages 18 and above provide their own consent and children ages 8-17 years give assent for study participation. Consent and assent documents are provided in English and translated to the local language (Luganda).

### Eligibility Criteria

Eligible participants include children and youth in Uganda, age 4-26 years with T1D (determined by clinical criteria) of at least 12 months duration, receiving insulin therapy, and having access to a cell phone (which is nearly ubiquitous in Uganda).

The exclusion criteria include those unwilling or unable to be seen monthly at the paediatric diabetes clinic, patients who are currently pregnant or breast-feeding, women likely to become pregnant in the next year, a major medical condition which the investigator feels would interfere with study participation, the patient already has CGM, inability to wear the sensor for at least 7 days or return it during the baseline assessment period, and the participant is deemed unlikely or unable to comply with the protocol.

### Study Management

Study participants will be seen monthly in clinic, where they will receive intensive monthly diabetes self-management education. Between clinic visits, they will have unlimited availability to contact study personnel by telephone. All participants must return used sensors at each clinic visit. Both groups will receive monthly education focused on pattern recognition and insulin adjustment to prevent hypo- and hyperglycemia.

Those participants wearing the unblinded FreeStyle Libre 2 CGM system will be able to see their glucose levels at all times. The research team will be able to use these data for insulin adjustment, and the participants will be taught how to interpret the data. Subjects in the control arm will wear the blinded CGM Libre Pro while performing SMBG 3x daily for 6 months. Blinded FreeStyle Libre Pro CGM sensors, placed monthly, will be used to provide control data. The study team will upload the data to a study website and neither participants nor the local research teams will have access to the blinded sensor data for clinical use until the end of months 0-6, to preserve the blinding.

### Study Progress

In August 2022, the Uganda study team was trained for 3 weeks on the protocol, CGM use, interpretation of both CGM and SMBG data, and research principles by the Minnesota study team. The study began recruitment in September 2022 and has enrolled 46 participants into the first of four planned 12-month cohorts. Video calls every 2-4 weeks and in-person visits by the Minnesota team 2-3 times per year will provide ongoing monitoring and mentorship throughout the study. Members of the Ugandan team have been invited to Minnesota for further training.

### Sample Size and Statistical Methods

#### Sample Size Determination

The sample size is determined based on hypothesis testing of the primary efficacy endpoint. The null hypothesis is that the two arms will demonstrate no significant differences in the 6- month change in glucose TIR. The alternative hypothesis is that wearing unblinded CGM will have greater improvement in glucose TIR over six months compared to the control arm. We estimate a standard deviation of 16.6% for baseline TIR based on our pilot data and assume a moderate correlation of 0.5 between baseline and 6-month TIR values. A sample size of 144 patients (72 per arm) will have over 90% power to detect a difference of 8.6% in 6-month TIR changes between the two arms at the one-sided significance level of 0.05. To allow a 20% dropout rate, a total of 180 patients will be randomized with planned study completion of at least 144 patients. The sample size will have over 90% power to detect a 5-percentage point difference in the 6-month change from baseline in percent time <70mg/dL (3.9 mmol/L) or a 3.5 percentage point difference in percent time < 54mg/dL (3.0 mmol/L) at the one-sided significance level of 0.05.

#### Analysis populations

The following populations will be considered for data summaries:

- The **enrolled population** will consist of all subjects who have signed an informed consent form and have begun the 2-week baseline evaluation procedure.
- The **intent-to-treat (ITT) population** is defined as all subjects in the enrolled set who have been randomized to one of the two treatment groups.
- The **per-protocol (PP) population** is defined as all subjects in the enrolled set who have been randomized and do not have major protocol deviations that may significantly impact the primary efficacy assessment.
- The **safety population** will consist of all subjects in the enrolled set who have received any treatment for glycemic control.

#### Primary Efficacy Analysis

The primary efficacy analyses will be based on the ITT population. Linear mixed effects models will be used to test the effects using unblinded CGM versus 3x/day SMBG (with blinded CGM to measure endpoint values) on the 6-month change rate of percent TIR, which adjust for subject-level effects and important covariates such as age and gender and account for auto-correlations among the repeated measures.

#### Secondary Efficacy Analysis

The primary endpoint will be also analyzed using the PP Population as a supportive analysis. The secondary endpoints, 12-month changes in time-in-range, time spent in other key glucose ranges (<70, <54, >180, >250 mg/dL) at 6 and 12 months, and 6- and 12-month changes in HbA1c will be analyzed using linear mixed effects models as for the primary endpoint. 95% confidence intervals and p-values will be calculated for the effects of using unblinded CGM on these endpoints. Analyses of patient satisfaction and quality of life will be descriptive. Summary statistics including mean, standard deviation, median, interquartile range, and range will be presented.

#### Cost analyses

In the primary cost analysis, the absolute cost of the Freestyle Libre 2 CGM system will be compared to that of SMBG 3x/day per person per 1 year. We will also calculate the cost of 6x/day SMBG per person per year. Secondary cost analyses include calculating the annual costs for each group for hospital admissions for severe hypoglycemia and DKA, and days missed from school or work. We will also estimate future long-term costs to the healthcare system based on published associations between time-in-range and risk of retinopathy and microalbuminuria.

Summary statistics will be calculated for each cost endpoint including mean, median, standard deviation, interquartile range, minimum and maximum by group. Their distributions will be plotted using side-by-side boxplots. The primary and secondary endpoints will be compared between the Freestyle Libre 2 CGM system and SMBG groups using 2-sample t-tests or the Wilcoxon rank-sum tests. Linear regression models will also be used for the group comparisons with adjustment for important covariates such as age, gender, and education levels.

#### Safety Analysis

Safety will be evaluated using the safety analysis set. The safety endpoints include episodes of severe (unconscious or requiring outside assistance regardless of sensor blood glucose level) hypoglycemia, DKA, superficial skin reactions, and overall study AEs and SAEs. The incidence and percentage of hypoglycemia, DKA, superficial skin reactions, overall study AEs and SAEs will be displayed for each treatment group. Severity and type of AEs, and relationship of AEs to study agent will also be summarized descriptively. Abnormal physical examination findings will be listed. No inferential statistical analyses are planned for safety endpoints.

#### Other Analysis

Descriptive analyses will be performed for the training endpoints.

## ETHICS AND DISSEMINATION

The protocol is approved by the Mulago Hospital Research Ethical Committee (MHREC 2173), Uganda National Council of Science and Technology (HS2129ES) and the University of Minnesota IRB (STUDY00013430). A Data and Safety Monitoring Board (DSMB) consists of 3 physicians, none of whom are otherwise associated with the study. The DSMB includes representatives from the US, Uganda, and Tanzania.

All study procedures will be performed by trained personnel, and all subjects will have phone access to local study personnel 24/7. CGM is considered a low-risk medical intervention. All participants will receive monthly education on how to prevent, recognize and treat both hyper- and hypoglycemia. For patients on the unblinded CGM, the sensor will provide continuous glucose levels, hypoglycemia/hyperglycemia alarms, and glucose trend arrows to help prevent hypoglycemia, which will offer them significantly more protection from this complication than they currently have. For those utilizing SMBG, we are providing 3 test strips a day. Patients currently average 2-3 strips per day. All participants will have access to unblinded CGM for at least half of the study period.

This study includes children because T1D is an important childhood disease associated with significant morbidity and mortality. The protocol offers the prospect of direct benefit including extensive diabetes education, increased blood glucose monitoring, and the potential, if the study hypotheses are correct, for improved diabetes control. The study is likely to yield general knowledge that is of importance for the understanding and treatment of T1D in children in less resourced settings.

The results will be disseminated by presentations at scientific meetings and policy briefings at the Ministry of Health, Uganda, and publication in peer reviewed journals.

## Data Availability

All data produced in the present study will be available upon reasonable request to the authors

## AUTHOR CONTRIBUTION

Study Conceptualization: TP-W and AM

Study Design: TP-W, CN, EP, EA, ETR, LZ, SB, AM

Study Performance: all authors

Manuscript preparation and editing: all authors

## PATIENT AND PUBLIC INVOLVEMENT STATEMENT

This study was preceded by a pilot study (4) to provide the baseline measures used in the sample size calculations. At the end of that study, subjects were enthusiastic about CGM and expressed the hope that it would become clinically available in the future to Ugandans. The involvement of Ministry of Health economists will help promote general availability if the current study is positive and if CGM proves to be cost effective. Throughout the study, questionnaires will be included that assess both the patient’s experience with diabetes and with CGM, and their assessment of the burden of research participation. The Minnesota and local Ugandan research teams will together lead at least one workshop directed at local patients and families which will cover the purpose of research, clinical research vs standard-of-care, the informed consent process, the concept of risk-benefit balance, and participant rights.

## REFERENCES

1. International Diabetes Federation. 2017. IDF Diabetes Altas. 8th edition.

2. Gregory GA, Robinson TIG, Linklater SE, Wang F, Colagivri S, de Beaufort C, Donaghue K, International Diabetes Federation Diabetes Atlas Type 1 Diabetes in Adults Special Interest Group, Magliano DJ, Maniam J, Orchard TJ, Rai P, Ogle GD. 2022. Global incidence, prevalence and mortality of type 1 diabetes in 2021 with projection to 2040: a modelling study. Lancet Diabetes Endocrinol 10: 741–60

3. Ogle GD, Kim H, Middlehurst AC, Silink M, Jenkins AJ. 2016. Financial costs for families of children with Type 1 diabetes in lower-income countries. Diabet Med 33: 820–6

4. McClure Yauch L, Velazquez E, Piloya-Were T, Wainaina Mungai L, Omar A, Moran A. 2020. Continuous glucose monitoring assessment of metabolic control in East African children and young adults with type 1 diabetes: A pilot and feasibility study. Endocrinol Diab Metab 00:e00135. doi.org/10.1002/edm2.135

5. Foster NC, Beck RW, Miller KM, Clements MA, Rickels MR, DiMeglio LA, Maahs D, Tambolane WV, Bergenstal R, Smith E, Olson BA, Garg S, TID Exchange Clinic Network. 2019. State of type 1 diabetes management and outcomes for the T1D Exchange in 2016-2018. Diabetes Technol Ther 21: 66–72

6. Miller KM, Foster NC, Beck RW, Bergenstal RM, DuBose SN, DiMeglio LA, Maahs DM, Tamborlane WV, for the T1D Exchange Clinic Network. 2015. Current state of type 1 diabetes treatment in the U.S.: updated data from the T1D Exchange clinic registry. Diabetes Care 38: 971–8

7. Miller KM, Beck RW, Bergenstal RM, Goland RS, Haller MJ, McGill JB, Rodriguez H, Simmons HH, Hirsch IR, T1D Exchange Clinic Network. 2013. Evidence of a strong association between frequency of self-monitoring of blood glucose and haemoglobin A1c levels in T1D Exchange Clinic Registry participants. Diabetes Care 36: 2009–14

8. DiMeglio LA, Acerini CL, Codner E, Craig ME, Hofer SE, Pillay K, Maahs D. 2018. ISPAD clinical practice guidelines 2018: Glycemic control targets and glucose monitoring for children, adolescents, and young adults with diabetes. Pediatr Diabetes 19 (Issue S27): 105–14

9. American Diabetes Association. 2023. Standards of Medical Care in Diabetes. Diabetes Care 46, suppl 1

10. Ogle GD, von Oettingen JE, Middlehurst AC, Hanas R, Orchard TJ. 2019. Levels of type 1 diabetes care in children and adolescents for countries at varying resource levels. Paediatric Diabetes 20: 93–8

11. Battelino T, Conget I, Olsen B, Schutz-Fuhrmann I, Hommel E, Hoogma R, Schlierloh U. N. S, Bolider J, and the SWITCH Study Group. 2012. The use and efficacy of continuous glucose monitoring in type 1 diabetes treated with insulin pump therapy: a randomized controlled trial. Diabetologia 55: 3155–62

12. Beck RW, Riddlesworth T, Ruedy K, Ahmann A, Bergenstal R, Haller S, Kollmah C, Kruger D, McGill JS, Polonsky W, Toschi E, Wolpert H, Price D, for the DIAMOND Study Group. 2017. Effect of continuous glucose monitoring on glycemic control in adults with type 1 diabetes using insulin injections. The DIAMOND Randomized Clinical Trial. JAMA 317: 371–8

13. Juvenile Diabetes Research Foundation Continuous Glucose Monitoring Study Group. 2008. Continuous glucose monitoring and intensive treatment of type 1 diabete. N Engl J Med 2008: 1464–76

14. Pickup JC, Freeman SC, Sutton AJ. 2011. Glycaemic control in type 1 diabetes during real time continuous glucose monitoring compared with self monitoring of blood glucose: meta-analysis of randomized controlled trials using individual patient data. BMJ 343:d3805

15. Tyndall V, Stimson RH, Zammitt NN, Ritchie SA, McKnight JA, Dover AR, Gibb FW. 2019. Marked improvement in HbA1c following commencement of flash glucose moniroing in people with type 1 diabetes. Diabetologia 62: 1349–56

16. Van Name MA, Kanapka LG, DiMeglio LA, Miller KM, Albanese-O’Neill A, Commissariat P, Corathers SD, Harrington KR, Hilliard ME, Anderson BJ, Kelley JC, Laffel LM, MacLeish SA, Nathan BM, Tamborlane WV, Wadwa RP, Willi SM, Williams KM, Wintergerst KA, Woerner S, Wong JC, DeSalvo DJ. 2022. SENCE Study, Long-term Continuous Glucose Monitor Use in Very Young Children With Type 1 Diabetes. J Diabetes Sci Technol 26: 10.1177/19322968221084667

17. Hellmund R, Weitgasser R, Blissett D. 2018. Cost calculation for a flash glucose monitoring system for UK adults with type 1 diabetes mellitus receiving intensive insulin treatment. Diabetes Research and Clinical Practice 138: 193–200

18. Shi L, Hellmund R. 2020. Cost comparison of flash continuous glucose monitoring with self-monitoring of blood glucose in adults with type 1 or type 2 diabetes using intensive insulin from a US private payer perspective. US Endocrinology, 2020;16: 24–30

